# Incidence of infectious diseases in children with *Mycobacterium tuberculosis* in Japan

**DOI:** 10.1101/2025.03.26.25324665

**Authors:** Yuko Hamaguchi, Takayuki Yamaguchi, Takashi Yoshiyama

**Affiliations:** The Research Institute of Tuberculosis, Tokyo, Japan; Shiga University, Shiga, Japan

## Abstract

**Background:** In Japan, the universal Bacillus Calmette–Guérin (BCG) immunization program for infants aged < 13 months remains in effect. Since 2005, the BCG vaccination history has been required to be reported, in addition to the clinical and treatment status of childhood TB, through a notification system and made available as epidemiological data.

**Methods:** To understand the epidemiology of childhood tuberculosis (TB) in Japan under the 2013 Bacillus Calmette–Guérin (BCG) immunization program modification in Japan, we performed descriptive analysis on the national surveillance data.

**Results:** The median percentage of annual vaccination coverage for infants aged <13 months was 97.0% during the observed period (2007–2022). The age at which the majority of infants received their vaccinations was 3 months before 2013 and 5 months since 2013.During the follow-up period, the number of childhood TB notifications and annual incidence decreased by approximately 60% and 40%, respectively. The analysis revealed correlation between childhood TB and both age and BCG vaccination history, while the impact of the 2013 BCG immunization program modification was not statistically significant.

**Conclusion:** These findings, derived from the national surveillance data in Japan, serve as a valuable resource for future research, offering insights into the efficacy of BCG and the impact of BCG policies.

## 1. Introduction

The Bacillus Calmette-Guérin (BCG) vaccine, which has been used for several decades to prevent tuberculosis (TB) caused by *Mycobacterium tuberculosis*, was developed in the early 20^th^ century. The vaccine contains a live and weakened strain of *Mycobacterium bovis*. Most research findings on the efficacy of BCG have been related to its protection against disease development, using observable data in TB cases. The efficacy of BCG in childhood TB is likely to be as high as 60–80% in preventing not only the development of TB but also severe diseases such as TB meningitis (1–3). Therefore, it is recommended that universal BCG immunization be initiated as early as possible during infancy, particularly in countries with high TB burden. This immunization program is designed to reduce the probability of newborn infants with susceptible immune systems becoming infected with TB and to prevent the progression to severe disease. Almost half a century has passed since most countries with low TB burden discontinued universal BCG vaccination. TB notification databases linked to BCG vaccination history are scarce in countries, including those in which universal or selective BCG vaccination has been introduced.

In Japan, a universal BCG immunization program for infants younger than 13 months is still ongoing. Since 2005, two modifications have been made to the BCG vaccination schedule for infants aged < 12 months (4). Children up to the age of 4 years were vaccinated until 2004. The initial and second modifications to the BCG immunization program were implemented in 2005 and 2013, respectively. The former involved establishing a vaccination schedule for infants up to six months of age (mostly between 3 and 5 months) to enhance the effectiveness of the BCG vaccine in preventing childhood TB. The latter involved prolonging the vaccination schedule up to 12 months of age (mostly between 5 and 7 months) due to an increase in reports of adverse reactions among infants who received the vaccine. BCG vaccination history is reported as childhood TB status through a reporting system and published as an epidemiological data. The objective of this study was to elucidate the dynamics underlying the incidence of childhood tuberculosis (TB) among individuals under the age of 5, and the potential ramifications of the policy modifications pertaining to the BCG vaccine.

## 2. Material and methods

### 2.1. Data collection

Data on BCG vaccinations for children aged 0–4 years with newly diagnosed pediatric TB (excluding latent TB infection) between 2005 and 2022 were obtained from the National Epidemiological Surveillance Information Database (NESID) (5). The NESID has collected BCG vaccination data since 2005, which are publicly accessible along with childhood TB notifications for children under five years of age.

Data on the number of births and annual BCG vaccination coverage among children born between 2005 and 2022 were obtained from the Ministry of Health, Labour and Welfare (MHLW) (6,7). We divided the aforementioned notification data from 2005 to 2022 into two datasets, one before and one after the 2013 BCG immunization program. These datasets were analyzed for our study.

### 2.2. Analysis

Descriptive epidemiological analysis was used as the primary analysis: the number of notifications of TB in children under 5 years of age was described, and the annual incidence rate (100,000 person-years) was calculated with its 95% confidence intervals, assuming a Poisson distribution. The Cochran–Armitage test was used to analyze the time trends in the above indicators. For these analyses, BCG immunization history was considered. Secondary analysis involved comparison of childhood TB incidence before and after the BCG policy change. To identify potential determinants of childhood TB incidence in an exploratory manner, a generalized linear model (GLM) was used, and adjustment was made for the periods before and after the 2013 vaccine policy change (as a dummy variable or having either the calendar year), vaccination history (dummy variable), and age.

### 2.3. Ethical approval

As the analyses in this study utilize data that are already of established academic value, widely used for research purposes, and publicly available no ethical review was required for this study.

## 3. Results

### 3.1. BCG vaccination coverage

Data on the cumulative BCG coverage from 2001 to 2022 are presented in Supplement 1. Overall, the median percentage of annual vaccination coverage for infants less than 12 months of age was 97.0% (interquartile range [IQR]: 94.0–97.7) for the follow-up period from 2007 to 2022. The median vaccination coverage for infants was 94.3% between 2007 and 2012 (the pre-2013 period), and the median vaccination coverage for infants was 97.7% between 2013 and 2022 (the post-2013 period).

In the pre-2013 period, most infants were vaccinated at approximately three months of age (with a cumulative vaccination coverage of 50%). In the post-2013 period, the majority of infants were vaccinated at approximately five months of age.

### 3.2. Childhood TB

#### 3.2.1. Notifications

Data on overall notifications (median) revealed a total of 28 cases (IQR: 23.3-31.5) throughout the study period. Additionally, there was a notable decline in the incidence of childhood TB, with a 60% reduction from 47 cases (95% CI:34.53-62.5) in 2007 to 21 cases (95% CI:13-32.1) in 2022 (Figure 1a).

**Figure 1.**
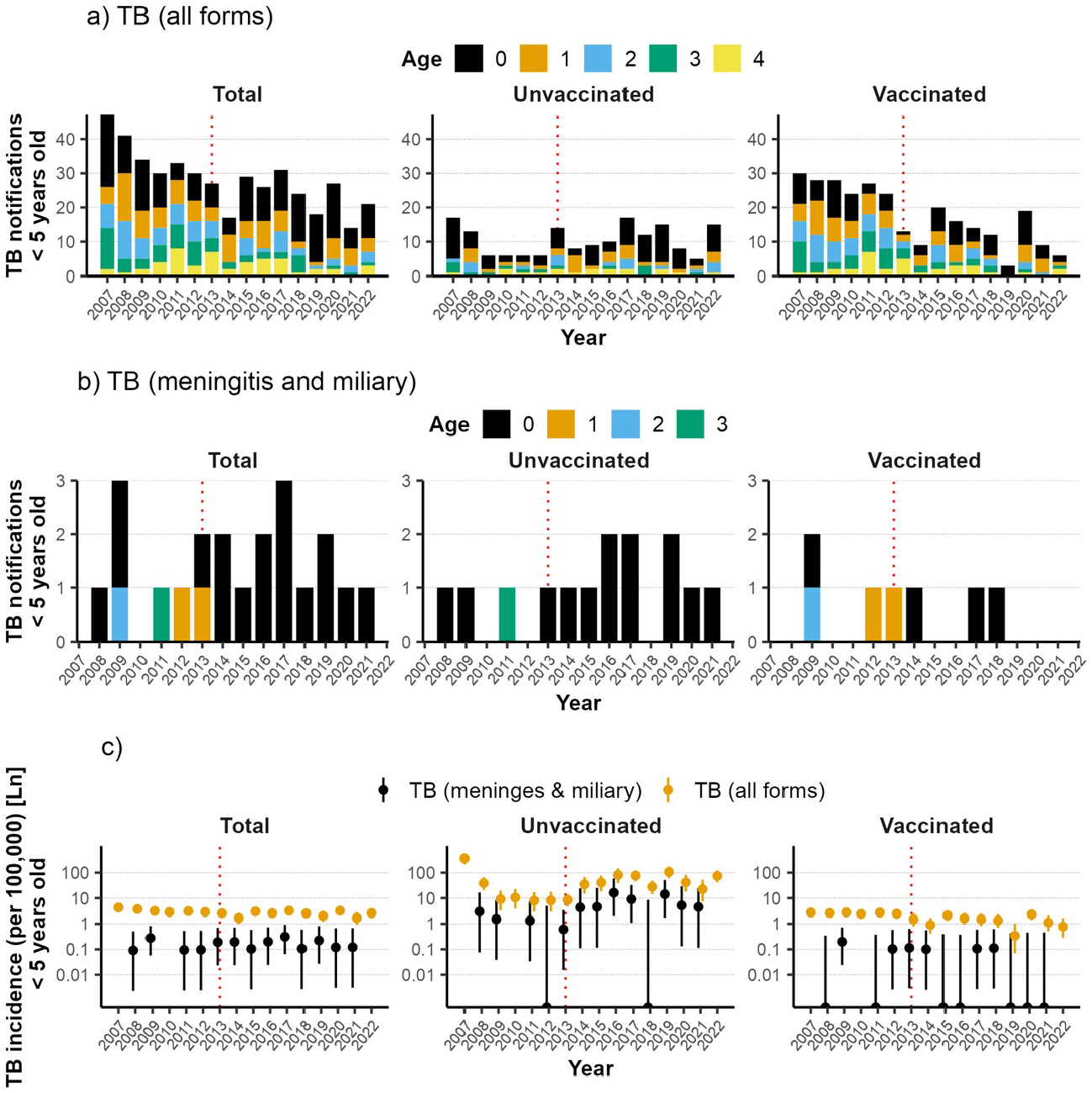
Childhood TB notifications and incidence between 2009 and 2022. The figure presents three distinct categories of data: a) annual notifications of all forms of tuberculosis (TB), b) notifications of severe cases (meningitis and miliary), and c) incidence rate per 100,000 person-years. The data are presented for children under five years of age, stratified by vaccination status. Furthermore, 95% confidence intervals (CIs) were calculated based on the assumption of a Poisson distribution.

While the median number of unvaccinated cases of TB was 9.5 (IQR: 6.0-14.3), exhibiting a fluctuating pattern between 17 and 5 cases, the number of vaccinated cases was 17.5 (IQR: 11.3– 24.8), demonstrating an 80% reduction from 30 cases in 2007 to 6 cases in 2022 (Figure 1a). Approximately half of these cases occurred in children aged 0–1 year.

The median number of notifications of severe TB in children exhibited a stagnating trend, with one case (IQR: 1.0–2.0) observed between 2007 and 2022 (Figure 1b). The respective numbers were 0.06 (IQR: 0.00–0.10) and 4.32 (IQR: 1.33–5.20) for vaccinated and unvaccinated children.

#### 3.2.2. Incidence

The TB notification rate (incidence) per 100,000 person-years among children was 2.87 (IQR: 2.58– 3.17) between 2007 and 2022. The incidence of childhood TB exhibited a notable decline, with an approximate 40% reduction from 4.30 (95% CI: 3.16–5.71) in 2007 to 2.63 (95% CI: 1.63– 4.02) in 2022. The most significant decline was observed at 1.67 (95% CI: 0.97–2.67) in 2014, following the BCG vaccination schedule modification (Figure 1c).

The incidence among vaccinated children was 1.87 (IQR: 1.28–2.51) over the course of the study period, representing a 20% decline from 2.75 (95% CI:1.86–3.93) in 2007 to 0.77 (95% CI: 0.28– 1.68) in 2022. The incidence among unvaccinated children exhibited considerable fluctuation, with a median of approximately 36.64 (IQR: 10.16–74.79) between 2009 and 2022.

The incidence of severe TB in children exhibited a relatively stable trend, with a median incidence of 0.12 (IQR: 0.11–0.20) between 2007 and 2022 (Figure 1c). Furthermore, the incidences of TB meningitis and miliary among the vaccinated and unvaccinated children reached a plateau, with median rates of 0.06 (IQR: 0.00–0.10) and 4.32 (IQR: 1.33–5.20), respectively.

### 3.3. Childhood TB by birth cohort

#### 3.3.1. Notifications

Birth cohort data were reformed to adjust for the impact of modifying the BCG vaccination schedule in the birth year rather than the notification data (Supplement 2). As the birth cohort data required included all ages from 0 to 4 years for analysis, the final dataset consisted of 12 birth cohorts between 2007 and 2018.

The data from the birth cohorts exhibited a trend similar to that observed in the notification data, with the majority of birth cohorts demonstrating the highest notifications among the age group 0 year, followed by 1 year, compared with other age groups (Figure 2a, Supplement 2). Analysis by country of birth showed few notifications, no increase or decrease, and no age-specific trends could be determined in the foreign born (Supplement 3).

**Figure 2.**
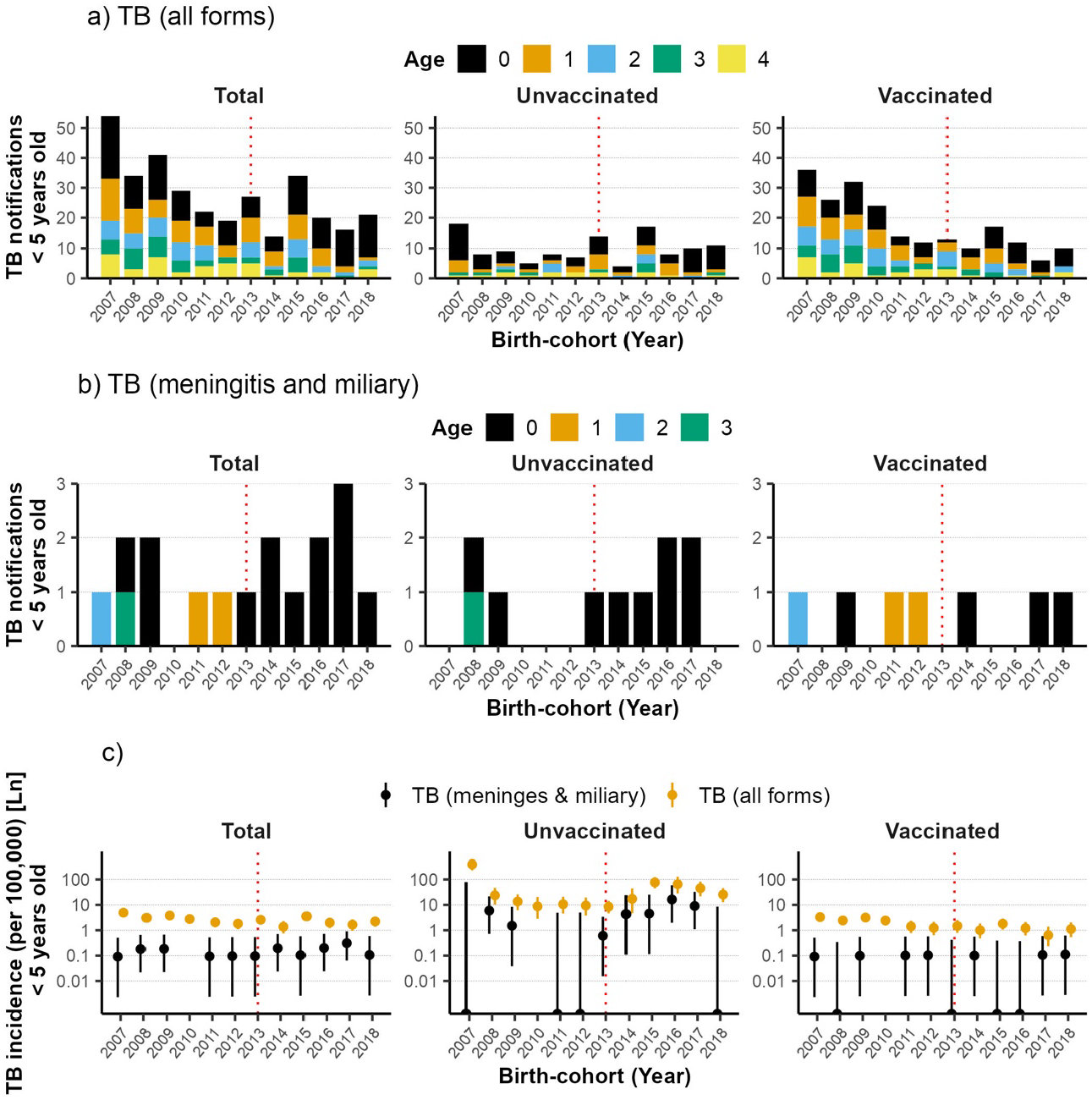
Annual notifications and incidence of childhood TB by birth cohorts between 2009 and 2018. The figure depicts three distinct categories of data: a) the annual notifications of all forms of tuberculosis (TB), b) the notifications of severe cases (meningitis and miliary), and c) the rate of incidence per 100,000 person-years. The data is presented for vaccinated and unvaccinated children under five years of age, organized by birth cohort. Additionally, 95% confidence intervals are calculated, assuming a Poisson distribution.

The 2007–2018 birth cohort data revealed 25 cases (IQR: 19.8–34.0) and exhibited a downward trend in notifications of childhood TB, with a 60% reduction from 54 cases (95% CI:40.57–70.46) born in 2007 to 21 cases (95% CI: 13–32.1) born in 2022 (Figure 2a). Following the alteration of the BCG vaccination schedule in 2013, there was a period of fluctuation in the number of TB notifications, with a notable decline in cases of children born in 2014 and a reduction of up to 14 cases. While the number of TB cases among the vaccinated cohort decreased by 30%, from 36 cases of children born in 2007 to 10 born in 2022, the number of cases among the unvaccinated cohort exhibited a stabilization at approximately nine cases (IQR: 7.8–11.8) of children born between 2007 and 2022.

The number of notifications of severe TB (meningitis and miliary) in children was one (IQR: 1–2) (Figure 2b).

The median number of notifications of TB meningitis and miliary in children demonstrated a stagnating trend in approximately one case (IQR: 1.0–2.0) observed between 2007 and 2018 (Figure 2b). The respective numbers were 0.06 (IQR: 0.00-0.10) and 4.32 (IQR: 1.33-5.20) for vaccinated and unvaccinated children.

#### 3.3.2. Incidence

The median notification rate (incidence) of TB meningitis and miliary TB in children born between 2007 and 2018 was 0.11 per 100,000 person-years (IQR: 0.09–0.19) (Figure 2c). The incidence of childhood TB stagnated from 0.09 (95% CI: 0–0.51) in children born in 2007 to 0.11 (95% CI: 0– 0.59) in children born in 2018.

The median incidence of TB meningitis and miliary disease among children born between 2007 and 2018 was 0.10 (IQR: 0.10**–**0.19) (Figure 2c). The incidence among vaccinated and unvaccinated children varied by median values of 0.10 (IQR: 0.00**–**0.10) and 1.51 (IQR: 0.00**–**5.24), respectively.

#### 3.3.3. Statistical analysis

Regarding childhood TB incidence before and after the vaccination schedule modification in 2013, a significant difference in the incidence of childhood TB was observed among both the vaccinated and unvaccinated birth cohorts (Welch two-sample t-test: p = 0.023). There were no significant differences between cohorts in terms of the incidences of TB meningitis and miliary disease.

Regarding the comparison of BCG vaccination history, a statistically significant but not notable difference in childhood TB incidence (Welch’s two-sample t-test: p = 0.095) was observed between the vaccinated and unvaccinated groups. For TB meningitis and miliary disease, there was a significant difference between the vaccinated and unvaccinated cohorts (Welch’s two-sample t-test: p = 0.034).

To identify the factors influencing childhood TB incidence in an exploratory manner, parameters were estimated in a generalized linear model (GLM), considering age, birth year, and BCG vaccination history (dummy variable with a value of 1 if vaccinated and 0 otherwise). The modification of the BCG vaccination schedule in 2013 was statistically significant when correlated with birth year (correlation coefficient: 0.889). Accordingly, the Akaike Information Criterion (AIC) was employed to ascertain the optimal model structure, using birth year as a variable. The analysis revealed that age and BCG vaccination history exhibited the most statistically significant and negative correlations (estimated coefficient: -0.82 and -0.97; p < 0.001 and = 0.029, respectively). The same approach was employed in the analysis of childhood meningitis and miliary resultant zone, where age and BCG vaccination history also demonstrated negative correlations (estimated coefficient: -0.50 and -0.94; p = 0.011 and 0.049, respectively).

## 4. Discussion

In this descriptive epidemiologic analysis conducted using TB notification data in children aged under 5 years, the median coverage rate of BCG mass immunization at 13 months of age was high (nearly 100%) throughout the observation period from 2007 to 2022. Slightly lower coverage was observed before 2013 than in the subsequent period. The objective of the 2013 BCG mass immunization modification was to postpone the immunization schedule for infants by approximately six months; however, the actual postponement in immunization was observed to be to 2–3 months, which was less than the intended duration.

A time-series analysis of birth cohort data by birth year between 2007 and 2018 revealed that the median number of notifications remained at 25 (IQR: 19.8–24.0) throughout this period. This represents a decrease of approximately 60% compared to previous years. The median annual incidence rate over 100,000 person-years was 2.41 (IQR: 1.95–3.20), representing a 50% decrease over the entire period. A comparison of the incidence of childhood TB between the pre-2013 and post-2013 periods revealed that except for BCG-vaccinated childhood TB cases, the incidence in the latter period was lower than that in the former. Furthermore, no significant discrepancy was observed between the incidence of BCG-vaccinated TB in children and BCG-unvaccinated TB in children.

The results of the multivariate-adjusted model analysis by GLM indicated that the incidence of childhood TB was associated with age and history of BCG vaccination. Similarly, GLM analysis revealed that the development of severe cases (TB meningitis and miliary disease) of childhood TB was associated with age and BCG vaccination history.

The results of the aforementioned analyses support the following conclusions: all forms of TB, including meningitis and miliary TB, are more prevalent in infants aged 0–1 year. This indicates that the risk of developing TB in this age group may be elevated owing to the immaturity of the immune system and that a history of BCG immunization is likely to be directly related to the risk of developing TB in this age group. Thus, it was inferred that the contribution of the 2013 modification of the BCG immunization program, which involved vaccinating infants 2-3 months after the pre-2013 period, to the increase in childhood TB was minimal.

The findings presented here are subject to certain limitations that must be considered when interpreting the results. The general recommendation is that infant BCG vaccination should be administered as early as possible after birth. However, the impact of a vaccination delay of only 2-3 months in vaccination on immune function in infants is currently uncertain. Furthermore, although there is some understanding of its impact on disease prevention, an exploratory descriptive analysis is insufficient to determine its impact on infection prevention. From April 2013 to March 2024, 377 cases of suspected serious adverse reactions to BCG vaccination were reported in Japan (8). Of these, 115 cases of BCG osteitis (osteomyelitis and periostitis), with a relatively clear causal relationship, were identified. This corresponds to an average annual incidence rate of 1.12 per 100,000 person-years. In comparison, the incidence rate of TB in children under five years of age was 0.11. Therefore, the number of cases of BCG osteitis was nearly ten times greater than that of childhood TB cases. Another report indicated that the average number of BCG osteitis cases per year was 1.3 before 2005, when BCG vaccination was initiated for children under 5 years old (9). However, from 2005 to 2012 and from 2013 to 2023, when BCG was administered to infants under six months of age, the average increased to 3.8 and 9.5 cases per year, respectively. This issue must be addressed with the utmost seriousness, particularly in severe cases. To implement an effective BCG policy, it is essential to develop a recommendation for BCG tailored to the present circumstances and based on the most recent evidence. It is hoped that further investigations and research will be conducted to utilize TB notification data in Japan with the aim of revealing and optimizing the efficacy of BCG in preventing infection and disease development in considering the relationship between side effects and the benefits of vaccination.

## 5. Conclusion

In conclusion, we shared the preliminary findings of a descriptive analysis of notification data on TB in children under the age of five years, including BCG vaccination history and treatment status of childhood TB. This epidemiological data on childhood TB in Japan represents a valuable resource for future research elucidating the efficacy of BCG as well as the impact of BCG policies.

## Supporting information

Spplementary Materials

## Data Availability

All data produced in the present study are available upon reasonable request to the authors

## Acknowledgements

This research was supported by the Japan Society for the Promotion of Science (JSPS; 20K18950 and 22K10417).

