## Supplementary material for "Incidence of infectious diseases in children with *Mycobacterium tuberculosis* in Japan": Spplementary Materials: Supplementary Materials medRxiv.pdf

**Supplement 1. Cumulative BCG vaccination coverage between 2001-2022**

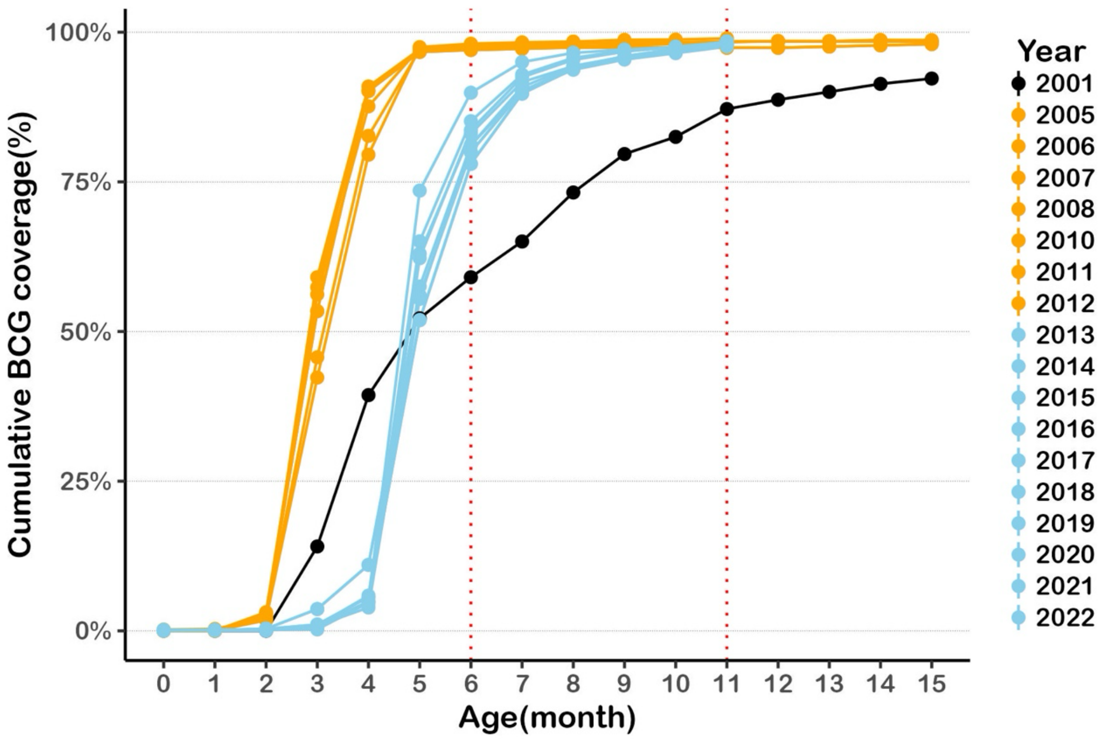

**Supplement 2. Age-specific childhood TB notifications by birth cohort between 2007-2018**

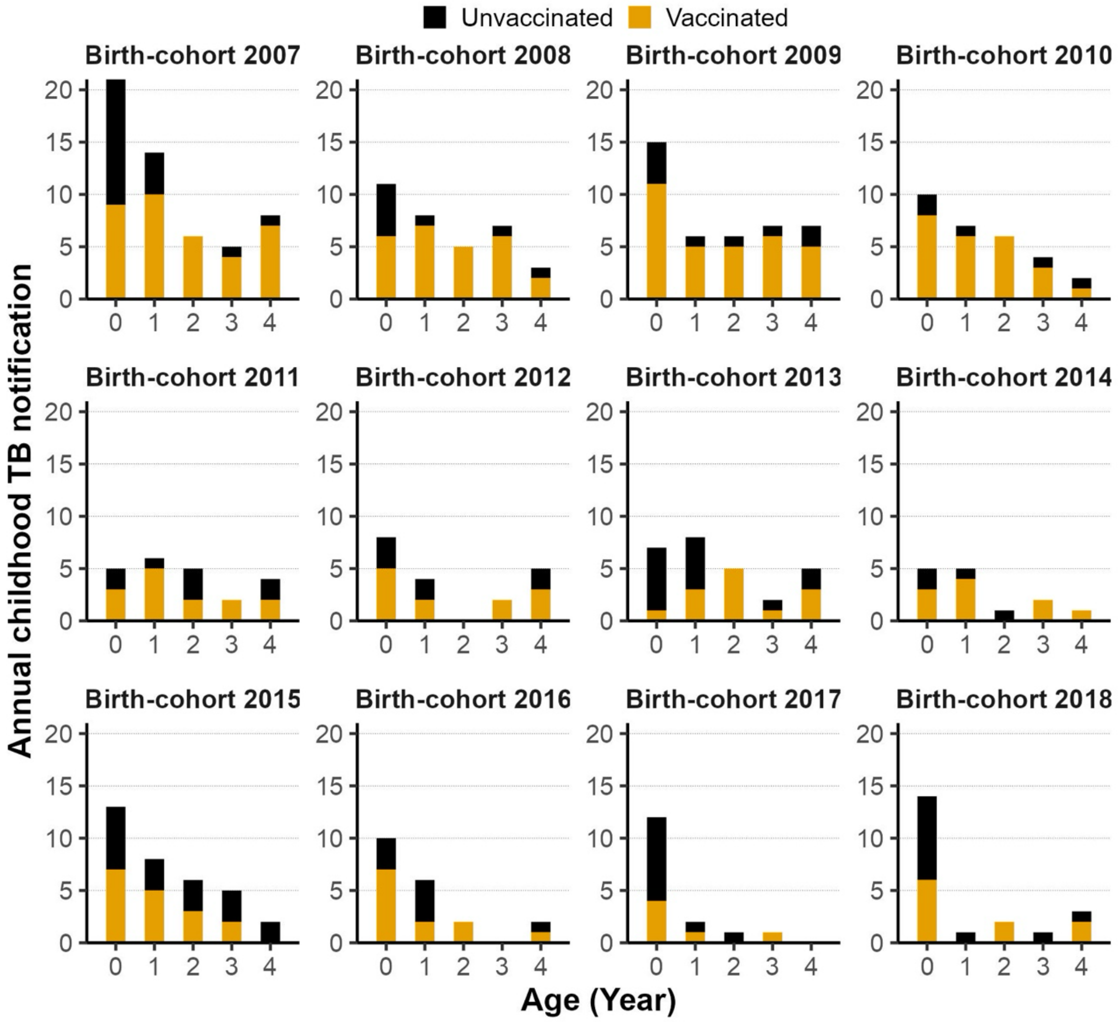

**Supplement 3. Age-specific childhood TB notifications in the foreign born by birth cohort between 2007 and 2018**

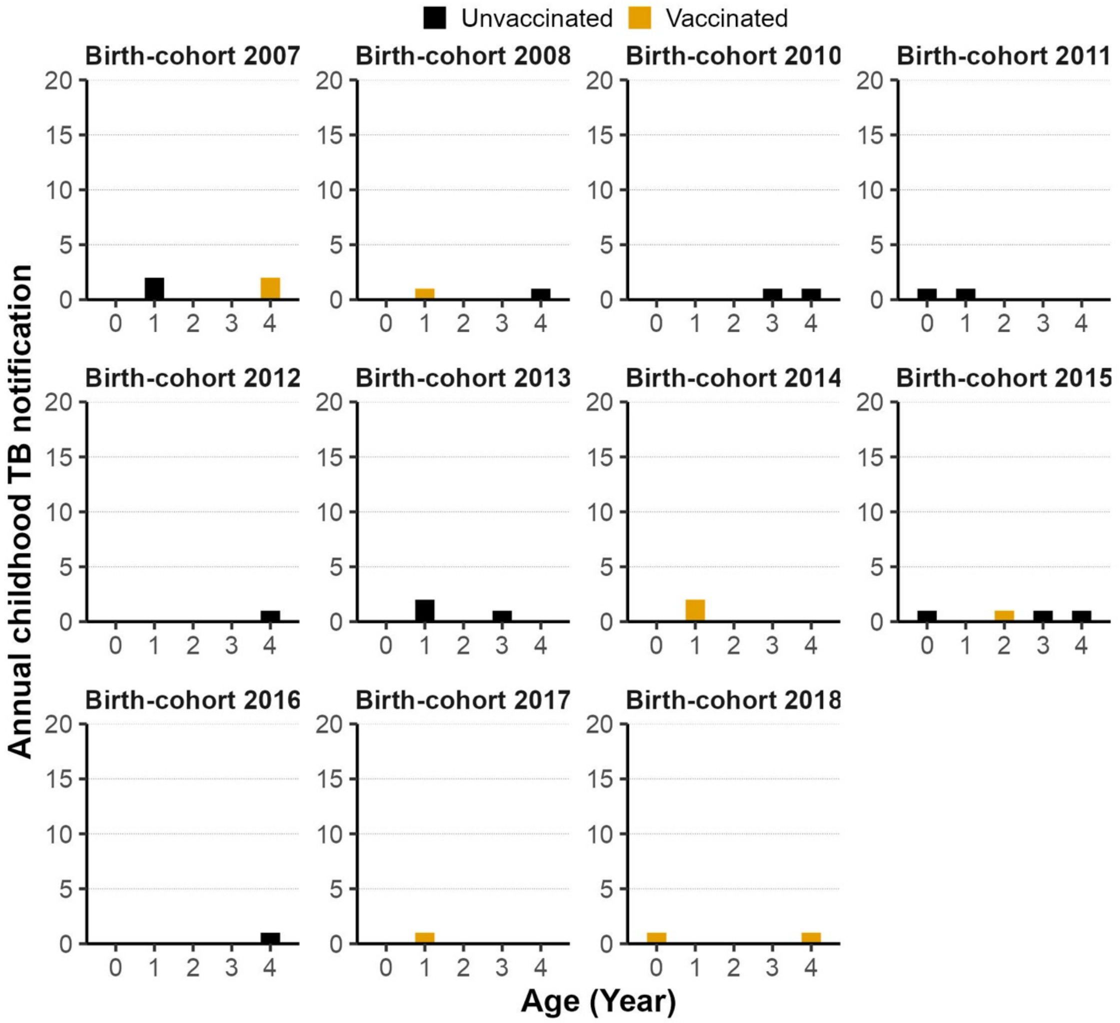
